# Understanding Food Insecurity in Kinshasa During the COVID-19 Pandemic

**DOI:** 10.1101/2024.03.06.24303901

**Authors:** Pierre Z. Akilimali, Benito Kazenza, Francis Kabasubabo, Landry Egbende, Steve Botomba, Dynah M. Kayembe, Branly K. Mbunga, Nguyen Toan Tran, Désiré K. Mashinda

**Affiliations:** Patrick Kayembe Research Center, Kinshasa School of Public Health, University of Kinshasa, Kinshasa, P.O. Box 11850, Democratic Republic of the Congo; (D.M.K.); (F.K.); Department of Nutrition, Kinshasa School of Public Health, University of Kinshasa, Kinshasa, P.O. Box 11850, Democratic Republic of the Congo; (L.E.); (B.K.); (S.B.); (B.J.M); Australian Centre for Public and Population Health Research, Faculty of Health, University of Technology Sydney, Sydney, P.O. Box 123, NSW 2007, Australia; Faculty of Medicine, University of Geneva, Rue Michel-Servet 1, 1206 Geneva, Switzerland; Department of Biostatistics and Epidemiology, Kinshasa School of Public Health, University of Kinshasa, Kinshasa, P.O. Box 11850, Democratic Republic of the Congo

**Keywords:** Slum areas, Food security, Kinshasa, Post-pandemic recovery

## Abstract

**Introduction:** Food insecurity is a vital issue, especially in places like Kinshasa. Additionally, food insecurity has been worsened by the COVID-19 pandemic, particularly in low- and middle-income countries. Thus, this study examined food insecurity in Kinshasa after the peak of the pandemic to understand food insecurity in post-pandemic recovery efforts and the possible implications for public health policies for future pandemics similar to COVID-19.

**Methods:** This study was conducted in Kinshasa with a representative sample of 2,160 households selected from 36 enumeration areas. We interviewed participants from different areas and used a questionnaire to ask them about their food situation. Interviews were conducted with the head of each household or their designated representative by 150 master’s students using tablets powered by the SurveyCTO application. Household food security status was evaluated through the Household Food Insecurity Access Scale. A logistic regression model was developed to assess household risk factors associated with food insecurity.

**Results:** Most people we talked to were over 40 years old, and many lived in households with fewer than six people. About a third of the households were overcrowded. Factors associated with food insecurity included being a household head aged over 50 years, insufficient living space, lower socioeconomic status, and residing in slum areas (AOR: 1.38; 95% CI: 1.06–1.79). In 2022, 12,627,424 individuals faced food insecurity in Kinshasa, including 8,829,820 individuals who experienced severe food insecurity.

**Conclusion:** Living conditions play a significant role in food insecurity. Governments need to do more to help people, especially those living in crowded areas. To combat economic restrictions that lead to food insecurity during crises, policymakers and implementing partners should enhance food assistance programs, such as cash transfers and food supply initiatives, focusing on overcrowded households and the informal job sector.

**Key Messages:** *What is already known on this topic:* This study emphasizes the multifaceted nature of food security, defined as the continuous access to sufficient, safe, and nutritious food, comprising availability, accessibility, utilization, and stability. Food insecurity, resulting from unmet needs in any of these dimensions, correlates with poor health outcomes and increased mortality. The global COVID-19 pandemic exacerbated food insecurity, particularly in low- and middle-income countries, with rates exceeding 50%. Factors such as poverty, living conditions, low income, lack of livestock, large household size, and psychological factors contribute significantly. While prior studies in the Democratic Republic of the Congo exist, they are limited, often focusing on specific groups. This study aims to comprehensively assess household food security in Kinshasa during the post-COVID-19 period, identifying associated factors for a more nuanced understanding.

*What this study adds:* This study adds to the existing literature by investigating the prevalence and determinants of food insecurity during a global health crisis, employing the Household Food Insecurity Access Scale for assessment. It contributes novel insights by examining the prevalence and severity of food insecurity in Kinshasa, the Democratic Republic of the Congo, offering a unique context for understanding the impact of a global health crisis on household food security.

*How this study might affect research, practice, or policy:* The study recommends implementing cash transfer strategies for vulnerable households, particularly those with informal jobs and young children, based on significant associations between lower socioeconomic status and food insecurity during the COVID-19 pandemic. Another recommendation is to expand food assistance programs for overcrowded households and the informal job sector, addressing the high prevalence of food insecurity in slum areas. Other social and structural determinants of food security, such as women’s empowerment and access to water and electricity, should be further researched.

## INTRODUCTION

Food security is achieved “when all people, at all times, have physical, social, and economic access to sufficient, safe, and nutritious food that meets their dietary needs and food preferences for an active and healthy life” [1–3]. This broad definition emphasizes four distinct dimensions of food security: availability, accessibility, and utilization of food, in addition to the stability of each of these factors, which refers to the ability to withstand shocks to the broader food system [1, 4]. Food insecurity occurs when at least one of these domain needs is not met, during which the experience at the household level may be temporary or longer [5–7]. Access to adequate food is a core social determinant of health, and food insecurity is related to poor nutritional intake and higher mortality rates [2, 6]. Even temporary reductions in food security can affect long-term health and cause a loss of human capital, from which it can take years to recover [8].

The recent COVID-19 pandemic has increased the level of food insecurity worldwide, and low- and middle-income countries have been most affected [8]. More than 50% of households experienced food insecurity during the COVID-19 pandemic [2, 6–8], and at the household level, it remains a major issue in many developing countries, particularly those in Africa. Although food insecurity remains high in low- and middle-income countries, many factors, such as poverty, exacerbate food insecurity. Food insecurity is also a significant risk factor for non-adherence to treatment such as antiretroviral therapy among HIV-infected individuals [9]. In contrast, factors such as living situation, low income, lack of livestock, high household size, and psychological situation (anxiety and depression) were principal aspects associated with food insecurity [2, 5, 8, 10].

Previous studies on food security have been conducted in the Democratic Republic of the Congo (DRC) over the past decade, but they have often been limited in scope, focused on specific demographic groups, or conducted on a relatively small scale. Further, these studies predate the COVID-19 pandemic and are often characterized by descriptive rather than analytical approaches. In the first half of 2020, Performance Monitoring for Action reported a 40% prevalence of food insecurity in the city of Kinshasa [11]. However, all urban residents do not uniformly experience food insecurity, and marginalized cities, commonly referred to as “slums,” represent the most significant examples of urban poverty in developing nations and are often the most impacted. Consequently, there is a pressing need to conduct a comprehensive assessment of food security on a broader scale, especially following the peak of the COVID-19 pandemic, and to delve into the factors contributing to food insecurity.

Slums are typically defined by homes that are hazardous, unhealthy, unstable, and overcrowded, without access to basic infrastructure and services. Slum residents in the city experience more health issues compared to non-slum residents [12]. As urban populations grow in developing countries, with the increasing population in informal urban areas in megacities worldwide, specific urban health intervention measures are crucial, particularly during times of crisis [12]. It is crucial to research and compare food insecurity levels between slum and non-slum residents, especially considering the health implications of urbanization and the potential exacerbation of these differences by events like the COVID-19 pandemic. The informal sector has grown vital for many individuals in the DRC due to the fast population expansion and a persistent shortage of legitimate job opportunities. Just 2.5% of employees work in the formal sector, and in 2021, the informal economy accounted for around 42% of the nation’s GDP [13]. The lockdown during the COVID-19 pandemic impacted workers in the informal sector, potentially affecting their food security.

Consequently, it is essential to comprehend food insecurity in post-pandemic recovery efforts and its possible implications for public health policies for future pandemics similar to COVID-19. Hence, the primary objective of this study is to gain insights into the status of food insecurity at the household level in Kinshasa during the COVID-19 pandemic and to identify the associated factors.

## METHODS

### Settings and conceptual framework

The DRC is the fourth most populous country in Africa [14]. Kinshasa, the capital of the DRC, is classified as one of the world’s “megacities.” In 2022, the metro area population of Kinshasa was comprised of 15,628,000 individuals [15]. Subsequently, Kinshasa ranks as Africa’s third-largest metropolis, following Lagos and Cairo, and is among the continent’s fastest-growing urban regions [14]. Kinshasa is segmented into 35 Health Zones, each of which is further divided into Health Areas. Insufficient access to water and sanitation, together with inadequate hygiene practices, malnutrition, and food insecurity, are identified as some of the primary risk factors contributing to mortality and disability in the country. The DRC reported its initial confirmed case of COVID-19 on March 10, 2020. As of September 3, 2022, there were a total of 92,942 cases, with 92,940 confirmed cases and 1,357 deaths, resulting in a case-fatality rate of 1.5% [16].

This study aligns with the conceptual framework of food security, which encompasses food availability influenced by crop production, livestock holdings, and market access; food access involving off-farm income and credit access; vulnerability to food shortages due to shock, livestock holding, and income levels; and utilization determined by age, sex, education level, and household size.

### Study design

We conducted a community-based cross-sectional study spanning July 27 to August 3, 2022. The survey had a two-stage cluster sampling design. Census enumeration areas (EAs) were randomly selected in the first stage using the National Statistical Institute sampling frame.

Data were collected in 36 EAs in Kinshasa. Each EA was divided into “segments” to streamline the field workers’ efforts. Each segment was intended to consist of approximately 500 households and be surveyed by a specific team. The number of segments in each EA was calculated by dividing the total number of households in the EA by the average segment size of 500 households. Within the selected EA or segment, a listing of all households was obtained and used to randomly select 60 households (second stage). In the selected households, the head of the household or their designated representative was interviewed on the day of the survey. In total, 2,160 households were sampled across 36 EAs, with 60 households chosen from the EAs or segments. Finally, this study included 2,020 households (response rate of 94%).

### Data collection

Data collection was conducted by 150 master’s students from the Kinshasa School of Public Health (KSPH) using tablets with the SurveyCTO program. They were taught research tools, ethics, and linguistic issues. The master’s students, who acted as interviewers, also received instruction on how to administer questionnaires during two training sessions conducted by the research team as part of their training. Interviews were conducted in Lingala, the local language in Kinshasa, or French. We employed reverse translation, with the assistance of bilingual academics, to assure linguistic and conceptual equivalency while translating things from French to Lingala. Information was gathered and examined anonymously. The survey questionnaire did not include any personal identities of the participants. The primary respondent at the household level was either the head of the household or their designated representative on the day of the survey. If a selected household was inaccessible or lacked a capable person to participate in the interview, interviewers were directed to make three separate visits at various times before considering the residence as absent or vacant.

KSPH teaching staff that act as supervisors played a vital role in overseeing the interviewers and maintaining data quality in the field. The collected data were routinely checked by supervisors before being transmitted to the server. This process occurred every evening during the data collection phase. Supervisors conducted quality control visits to verify the correctness and completeness of data. It was a method of verifying if the master’s student had interviewed the appropriate respondents effectively. Quality control visits were conducted on 5% of the households in each EA. The quality check was conducted using a brief questionnaire that solely contained questions from the Household Food Insecurity Access Scale (HFIAS).

The master’s students were trained to verify the completeness and quality of their work, and the supervisors reviewed all data forms before their submissions. Forms containing omissions and clear errors were sent back to the master’s student through their supervisor for rectification or further review. The forms were checked for errors or inconsistencies at the time of data entry. Additionally, the central team conducted daily data visualization using the SurveyCTO server to provide timely feedback throughout the data collection period.

### Measurements

#### Sociodemographic variables

The sociodemographic variables included age, sex, ethnicity, size of household, annual household income, relationship status, employment status at the time of the survey, and whether the housing was located in an informal settlement (slums) [17]. Overcrowded households were defined as those with four or more persons living in one room. Conversely, a household was deemed to have sufficient living space when three or fewer people were living in one room.

#### Food security

Household food security status was estimated using the HFIAS [18], which is a validated instrument that distinguishes food-insecure households from food-secure households across different cultural contexts. The Nutrition Risk Screening 2002 tool comprises nine questions designed to reflect the following universal domains of the experience of food insecurity: (1) anxiety and uncertainty about household food supply, (2) insufficient quality (including variety and preferences for the type of food consumed), and (3) insufficient food intake and its physical consequences. The current results are presented in the following categorical format: (1) food secure, (2) mildly food insecure, (3) moderately food insecure, and (4) severely food insecure [19]; we dichotomized these categories into food insecure versus food secure. The dependent variable (household food security status) was a dichotomous variable that was assigned a value of 1 if the household was food secure and 0 otherwise. The Cronbach’s alpha was 0.94, demonstrating excellent internal consistency. The HFIAS has been utilized in prior research in the DRC [9]. Our study used the version that was previously translated into Lingala and the three other local languages.

While household food insecurity estimates provide an overall picture of a household’s access to food, they may not accurately reflect individual experiences within the household. Factors such as intra-household distribution of resources and varying dietary needs can impact individual food security. To gain a more nuanced understanding, combining household-level data with individual-level information or conducting separate assessments for individuals within the household may be more informative.

### Data analysis

All statistical analyses were conducted using STATA 17 (StataCorp, College Station, TX). Initially, we determined an overview of participants’ sociodemographic characteristics, both in their entirety and categorized by food security status. This involved employing cross-tabulations and chi-square tests to identify significant differences between non-slum and slum households. Significant differences in food security status were evaluated using chi-square tests. To assess household susceptibility to food insecurity as the primary outcome variable, a multivariable logistic regression model was developed. Factors associated with food insecurity in bivariate analysis were entered into a logistic regression model to obtain adjusted odds ratios (AOR) and their 95% confidence intervals (95%CI). To assess how the association between slum neighborhood and food insecurity might differ by living space status, an interaction term between living space status and slum residence was included in the multivariable model, and the log-likelihood ratio test was used to assess its significance.

The Breslow-day test for assessing the interaction effect was used. If it was found to be significant at *p* < 0.05, separate multivariate regression analyses were performed by type of neighborhood. The interactions between living space status and slum residence, between living space status and wealth index, and between wealth index and slum residence did not suggest heterogeneity between slum and non-slum residents. All the statistical analyses were conducted using Stata Version 17.0. SVY procedures in Stata and were used to account for the sampling design and selection weights. ORs and 95% CIs were estimated from the regression parameters. Variance-inflation factors were calculated to test for multicollinearity, with the highest found to be 2.65. The significance level was set at p< 0.05.

### Ethical considerations

The study was conducted in accordance with the principles outlined in the Helsinki Declaration. The master’s students were trained to obtain informed consent. Respondents who could not express themselves in French were offered consent forms in their preferred language. Ethical clearance was obtained from the ethical review board of the KSPH (no. ESP/CE/71B/2022). Each participant had the informed consent form read aloud to them and provided verbal consent. To standardize the informed consent process for illiterate participants, we opted for verbal consent witnessed by a third party. The third party ensured that the consent was read to the subject, who then freely agreed to participate in the study.

The participant was provided with a signed copy of the consent form to retain. The consent acquisition process was sanctioned by the ethics committee. This study did not involve any individuals under the age of 18. Information was gathered and examined anonymously. The survey form did not include any personal identifying of participants, and the participants were notified that their involvement was optional. They have the liberty to either accept, decline participation, or withdraw at any time without facing any consequences.

## RESULTS

### Participants’ characteristics

Overall, most individuals identified as the head of the household and were at least 40 years old (72%). Almost three-quarters of the participants were men (73.6%) and married (72.5%), and 60% had a schooling level between secondary school and university. Table 1 illustrates that the mean household size was six persons or fewer (75%), approximately one-third of the households were overcrowded (29%), and the socioeconomic status (SES) distribution among households was not equal. For these characteristics, the differences between people living in non-slums and people living in slums were statistically significant.

**Table 1.**
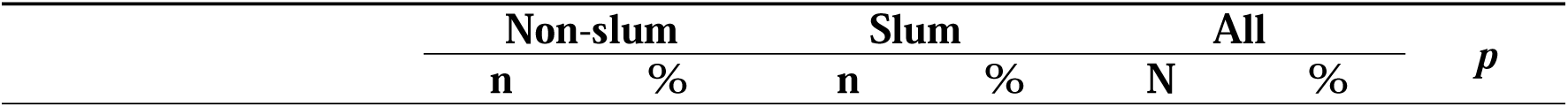
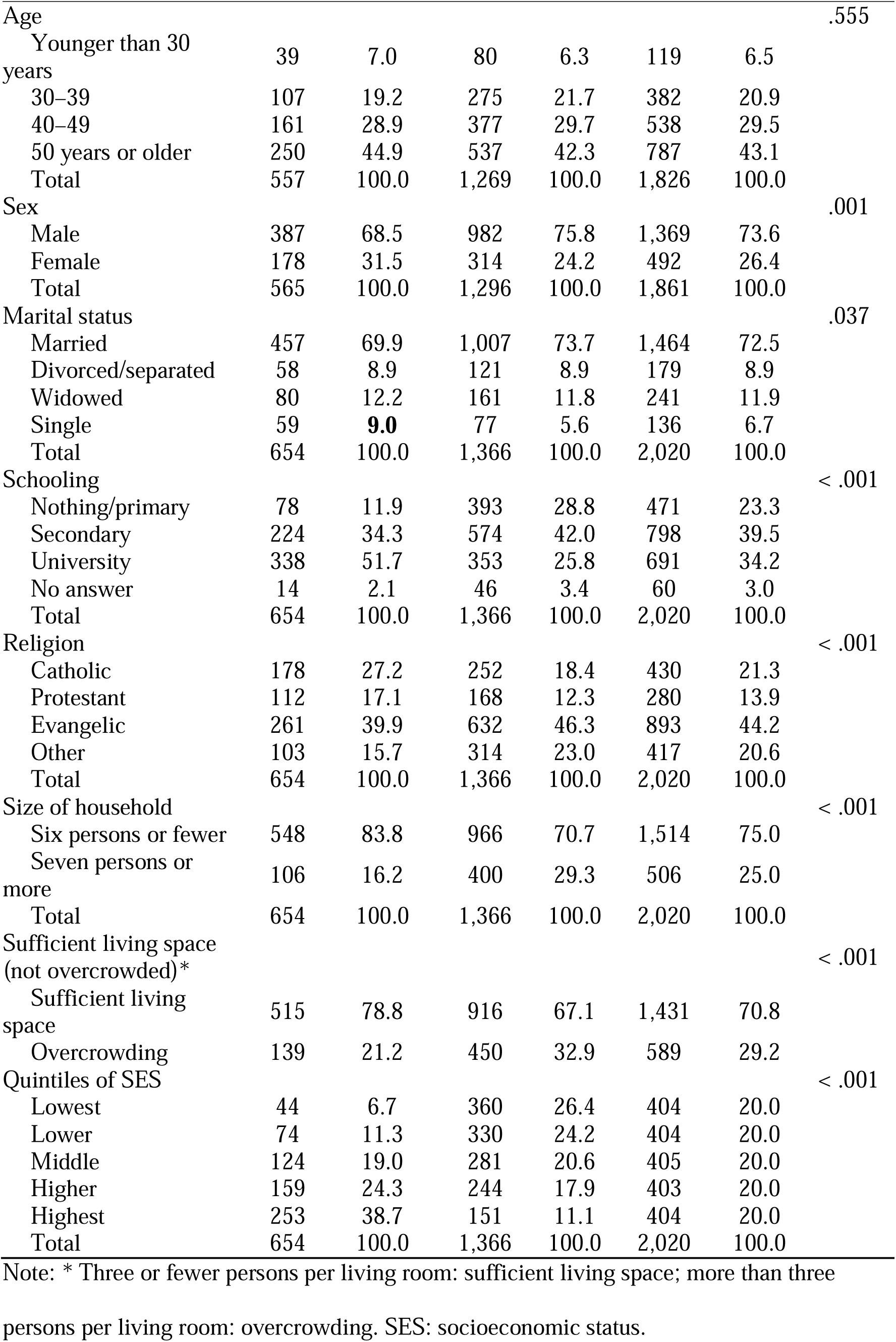
Sociodemographic characteristics of the head of the household.

| Non-slum |  | Slum |  | All |  | *p* |
| --- | --- | --- | --- | --- | --- | --- |
| **n** | **%** | **n** | **%** | **N** | **%** |  |

|  |  |  |  |  |  |  |  |
| --- | --- | --- | --- | --- | --- | --- | --- |
| Age |  |  |  |  |  |  | .555 |
| Younger than 30 years | 39 | 7.0 | 80 | 6.3 | 119 | 6.5 |  |
| 30–39 | 107 | 19.2 | 275 | 21.7 | 382 | 20.9 |  |
| 40–49 | 161 | 28.9 | 377 | 29.7 | 538 | 29.5 |  |
| 50 years or older | 250 | 44.9 | 537 | 42.3 | 787 | 43.1 |  |
| Total | 557 | 100.0 | 1,269 | 100.0 | 1,826 | 100.0 |  |
| Sex |  |  |  |  |  |  | .001 |
| Male | 387 | 68.5 | 982 | 75.8 | 1,369 | 73.6 |  |
| Female | 178 | 31.5 | 314 | 24.2 | 492 | 26.4 |  |
| Total | 565 | 100.0 | 1,296 | 100.0 | 1,861 | 100.0 |  |
| Marital status |  |  |  |  |  |  | .037 |
| Married | 457 | 69.9 | 1,007 | 73.7 | 1,464 | 72.5 |  |
| Divorced/separated | 58 | 8.9 | 121 | 8.9 | 179 | 8.9 |  |
| Widowed | 80 | 12.2 | 161 | 11.8 | 241 | 11.9 |  |
| Single | 59 | **9.0** | 77 | 5.6 | 136 | 6.7 |  |
| Total | 654 | 100.0 | 1,366 | 100.0 | 2,020 | 100.0 |  |
| Schooling |  |  |  |  |  |  | < .001 |
| Nothing/primary | 78 | 11.9 | 393 | 28.8 | 471 | 23.3 |  |
| Secondary | 224 | 34.3 | 574 | 42.0 | 798 | 39.5 |  |
| University | 338 | 51.7 | 353 | 25.8 | 691 | 34.2 |  |
| No answer | 14 | 2.1 | 46 | 3.4 | 60 | 3.0 |  |
| Total | 654 | 100.0 | 1,366 | 100.0 | 2,020 | 100.0 |  |
| Religion |  |  |  |  |  |  | < .001 |
| Catholic | 178 | 27.2 | 252 | 18.4 | 430 | 21.3 |  |
| Protestant | 112 | 17.1 | 168 | 12.3 | 280 | 13.9 |  |
| Evangelic | 261 | 39.9 | 632 | 46.3 | 893 | 44.2 |  |
| Other | 103 | 15.7 | 314 | 23.0 | 417 | 20.6 |  |
| Total | 654 | 100.0 | 1,366 | 100.0 | 2,020 | 100.0 |  |
| Size of household |  |  |  |  |  |  | < .001 |
| Six persons or fewer | 548 | 83.8 | 966 | 70.7 | 1,514 | 75.0 |  |
| Seven persons or more | 106 | 16.2 | 400 | 29.3 | 506 | 25.0 |  |
| Total | 654 | 100.0 | 1,366 | 100.0 | 2,020 | 100.0 |  |
| Sufficient living space (not overcrowded)* |  |  |  |  |  |  | < .001 |
| Sufficient living space | 515 | 78.8 | 916 | 67.1 | 1,431 | 70.8 |  |
| Overcrowding | 139 | 21.2 | 450 | 32.9 | 589 | 29.2 |  |
| Quintiles of SES |  |  |  |  |  |  | < .001 |
| Lowest | 44 | 6.7 | 360 | 26.4 | 404 | 20.0 |  |
| Lower | 74 | 11.3 | 330 | 24.2 | 404 | 20.0 |  |
| Middle | 124 | 19.0 | 281 | 20.6 | 405 | 20.0 |  |
| Higher | 159 | 24.3 | 244 | 17.9 | 403 | 20.0 |  |
| Highest | 253 | 38.7 | 151 | 11.1 | 404 | 20.0 |  |
| Total | 654 | 100.0 | 1,366 | 100.0 | 2,020 | 100.0 |  |

Note: \* Three or fewer persons per living room: sufficient living space; more than three
persons per living room: overcrowding. SES: socioeconomic status.

### Food security conditions of the household

Significant differences were observed between non-slum and slum households across all food security measurement items. More than half the households expressed concerns related to food scarcity, with 66.2% of all households reporting apprehensions about preferred food availability (non-slum: 59.0, slum: 69.6; *p* < .001). Additionally, 63.7% of the households reported that a lack of resources prevented household members from consuming their preferred types of food (non-slum: 52.0, slum: 69.3; *p* < .001). Further, in less than half of the households, there were instances in which individuals either went to bed hungry (31.0%) or had to endure an entire day without eating owing to insufficient food (50.4%; Table 2).

**Table 2.**
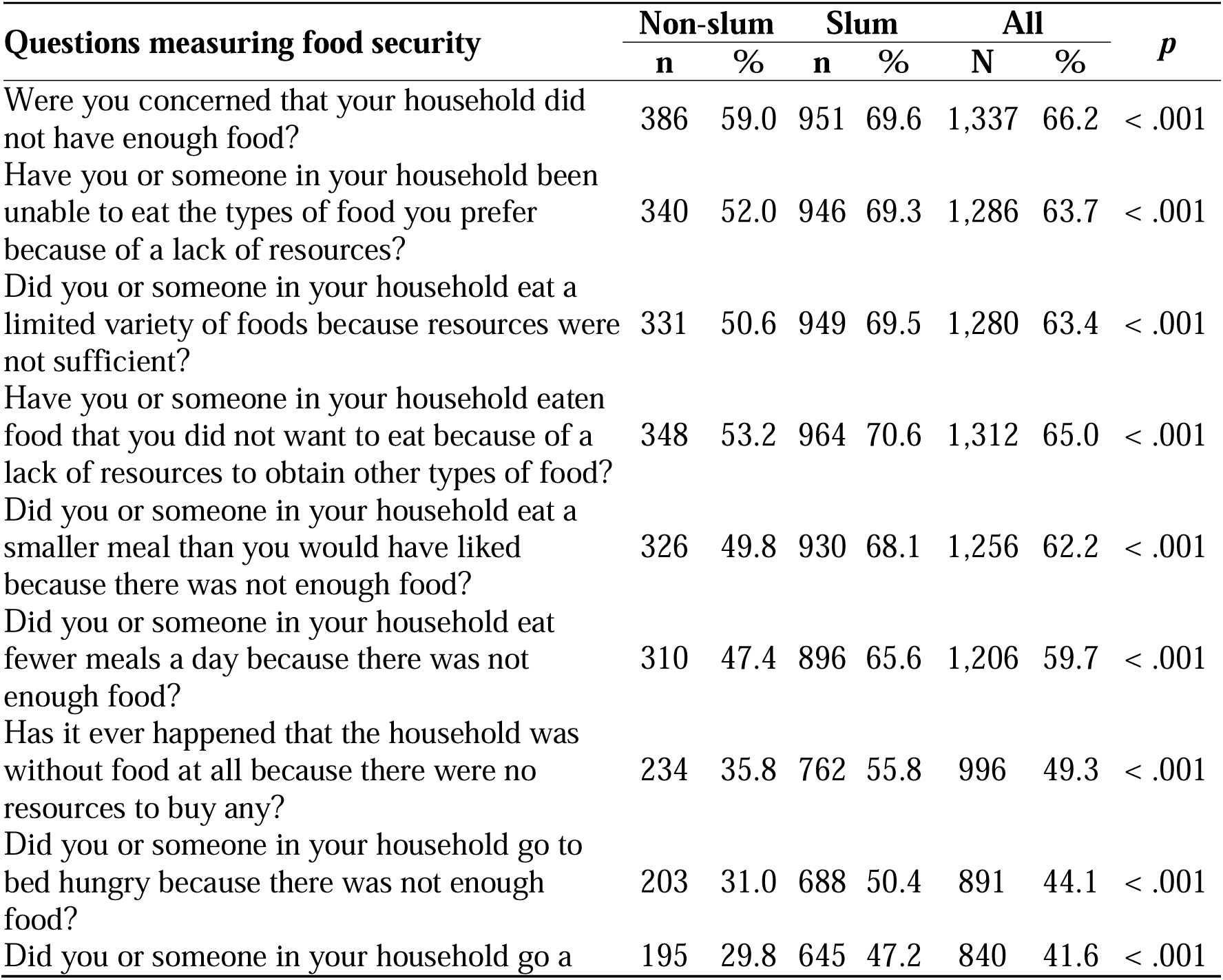
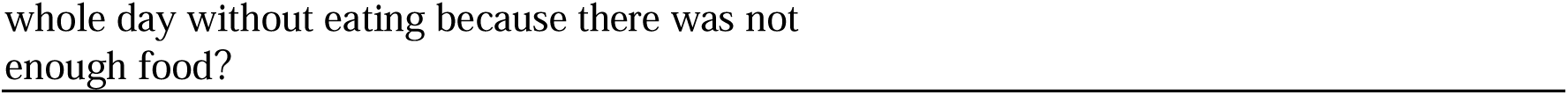
Households by residence and components of the nine food security conditions.

### Food security conditions of the individuals

Table 3 illustrates that during the COVID-19 pandemic, a similar pattern emerged for individuals residing in both non-slum and slum areas. Approximately 10% of the participants expressed concerns regarding insufficient food availability, limited access to preferred foods, dietary variety, or having fewer meals. Additionally, fewer than 5% experienced the hardship of going to bed hungry or enduring an entire day without food. Further, in at least 5% of the households, a complete absence of food was reported, and the absence was primarily attributable to resource constraints.

**Table 3.**
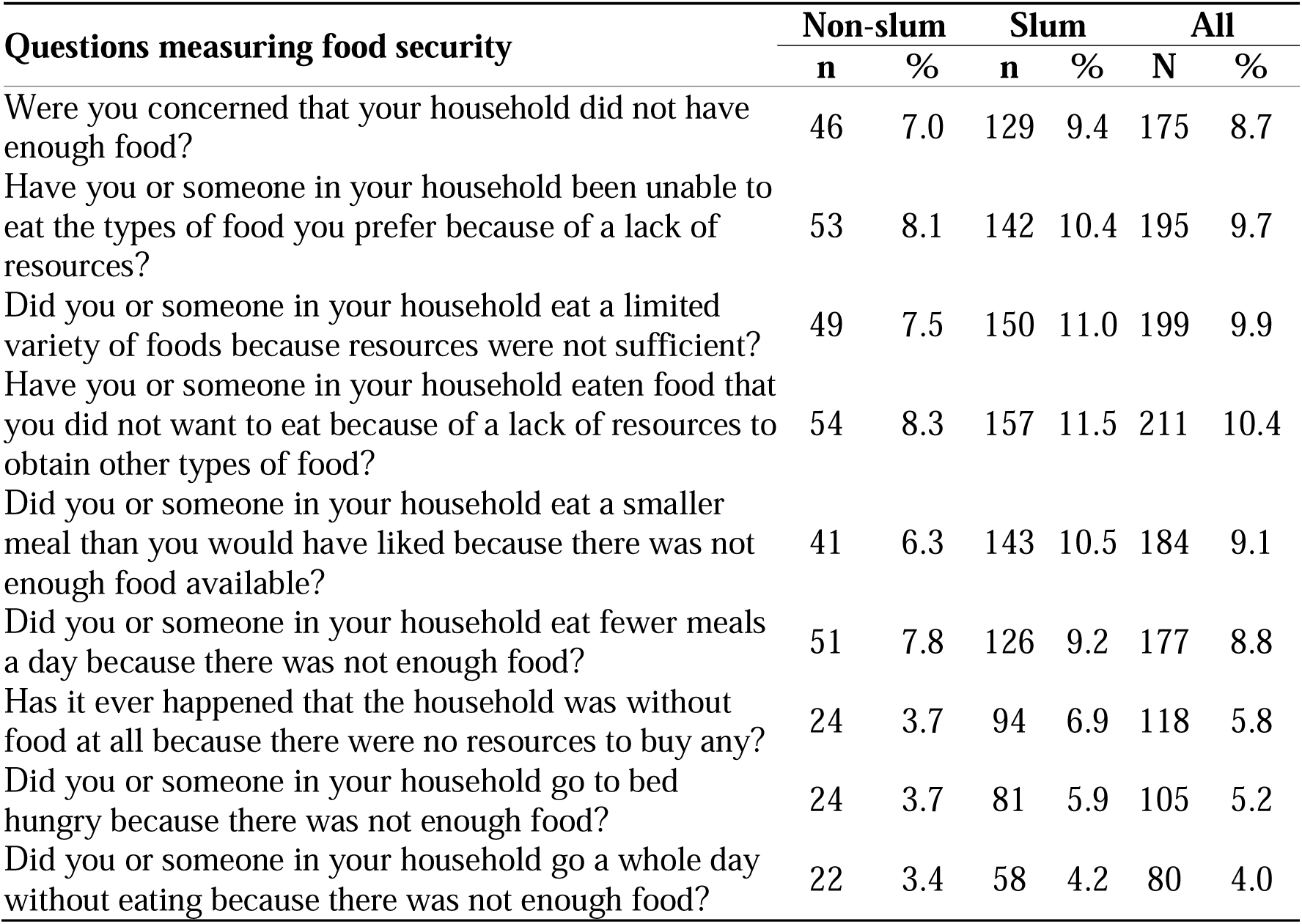
Food security conditions.

| Questions measuring food security | Non-slum |  | Slum |  | All |  |
| --- | --- | --- | --- | --- | --- | --- |
|  | n | % | n | % | N | % |
| Were you concerned that your household did not have enough food? | 46 | 7.0 | 129 | 9.4 | 175 | 8.7 |
| Have you or someone in your household been unable to eat the types of food you prefer because of a lack of resources? | 53 | 8.1 | 142 | 10.4 | 195 | 9.7 |
| Did you or someone in your household eat a limited variety of foods because resources were not sufficient? | 49 | 7.5 | 150 | 11.0 | 199 | 9.9 |
| Have you or someone in your household eaten food that you did not want to eat because of a lack of resources to obtain other types of food? | 54 | 8.3 | 157 | 11.5 | 211 | 10.4 |
| Did you or someone in your household eat a smaller meal than you would have liked because there was not enough food available? | 41 | 6.3 | 143 | 10.5 | 184 | 9.1 |
| Did you or someone in your household eat fewer meals a day because there was not enough food? | 51 | 7.8 | 126 | 9.2 | 177 | 8.8 |
| Has it ever happened that the household was without food at all because there were no resources to buy any? | 24 | 3.7 | 94 | 6.9 | 118 | 5.8 |
| Did you or someone in your household go to bed hungry because there was not enough food? | 24 | 3.7 | 81 | 5.9 | 105 | 5.2 |
| Did you or someone in your household go a whole day without eating because there was not enough food? | 22 | 3.4 | 58 | 4.2 | 80 | 4.0 |

### Level of food insecurity

Table 4 reveals that only 19.2% of participants were classified as food secure, leaving 80.8% (100%–19.2%) as food insecure, with 56.5% experiencing severe food insecurity.

**Table 4.**
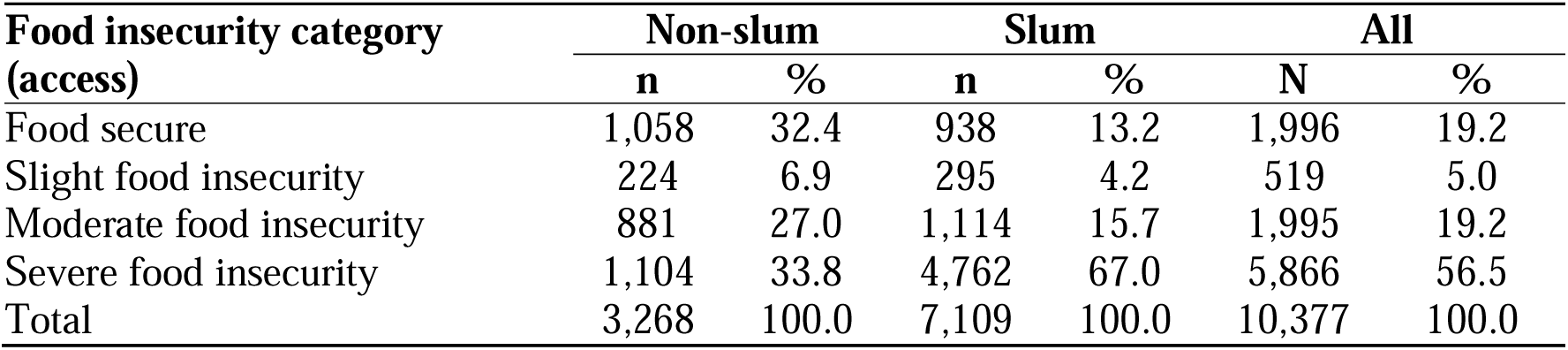
Distribution of food insecurity in the total population by accessibility pillar.

Extrapolating these percentages to the estimated population of 15,628,000 in Kinshasa, 12,627,424 individuals (0.808 * 15,628,000) were deemed food insecure in 2022, including 8,829,820 individuals experiencing it severely (0.565 * 15,628,000).

Notably, a marked distinction emerges when comparing residents of non-slum and slum areas within our sample in terms of severe food insecurity. Among respondents not residing in slums, around a third (33.8%) experienced severe food insecurity, contrasting sharply with approximately two-thirds (67.9%) of those living in slums.

### Factors associated with food insecurity

Figure 1 illustrates the factors associated with food insecurity. Specifically, individuals aged 50 years or older exhibited a significantly greater likelihood of experiencing food insecurity (AOR: 2.02; 95% confidence interval (CI): 1.23–3.31) as compared to their counterparts.

**Figure 1.**
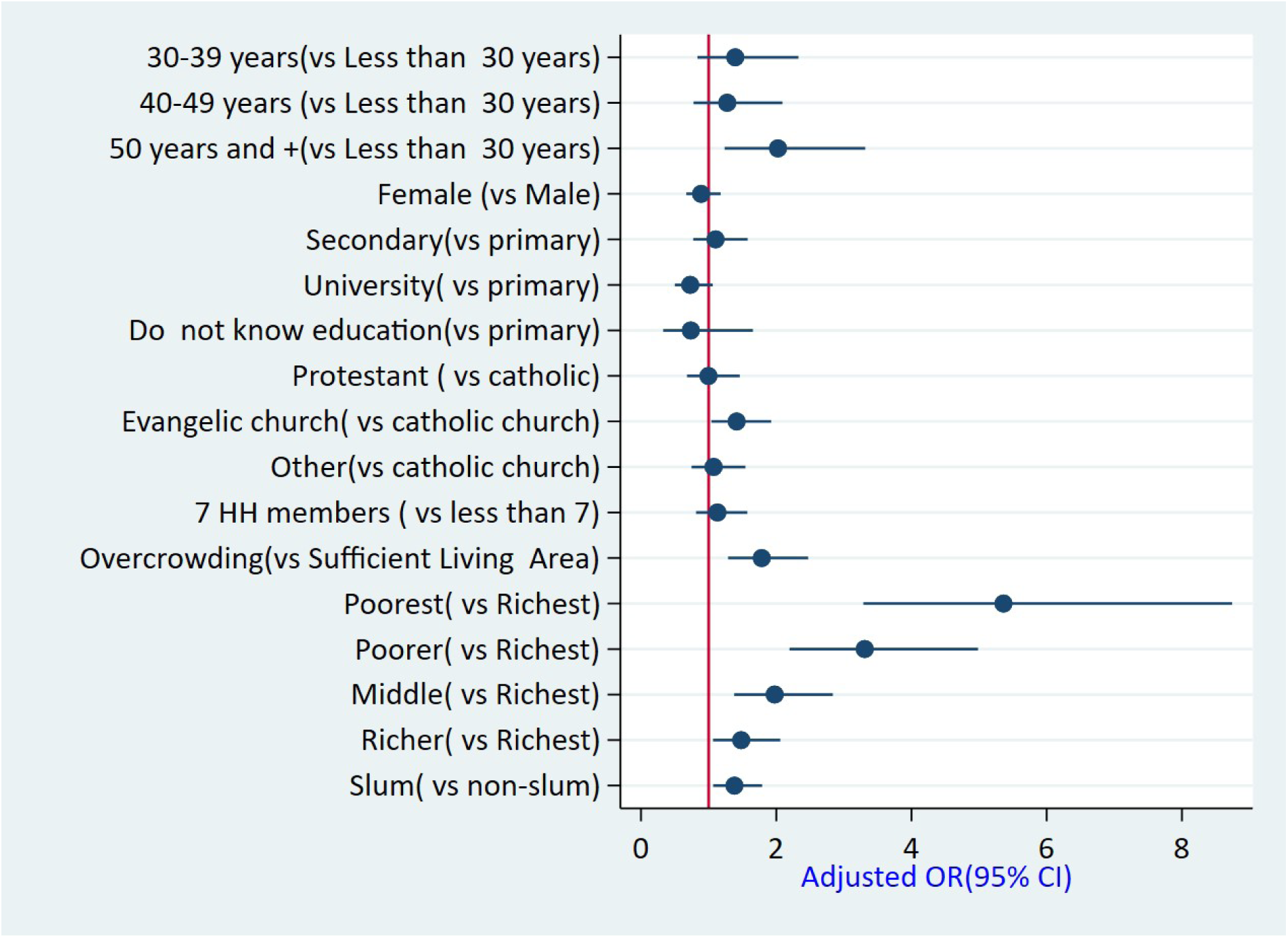
Forecast plot presenting the associated factors of food insecurity with their adjusted ORs 30-39 years(vs Less than 30 years) 40-49 years (vs Less than 30 years) SO years and +(vs Less than 30 years) Female (vs Male) Secondary(vs primary) University( vs primary) Do not know education(vs primary) Protestant ( vs catholic) Evangelic church( vs catholic church) Other(vs catholic church) 7 HH members ( vs less than 7) Overcrowding(vs Sufficient Living Area) Poorest( vs Richest) Poorer( vs Richest) Middle( vs Richest) Richer( vs Richest) Slum( vs non-slum)

Conversely, having a sufficient living space was associated with a reduced likelihood of food insecurity (AOR: 0.56; 95% CI: 0.44–0.77). Additionally, households characterized by lower SES (AOR lowest: 5.36; 95% CI: 3.29–8.74; AOR low: 3.30; 95% CI: 2.19–4.98; AOR moderate: 1.97; 95% CI: 1.37–2.84; AOR higher: 1.48; 95% CI: 1.06–2.06) and residing in slum areas displayed a heightened risk of food insecurity (AOR: 1.38; 95% CI: 1.06–1.79) as compared to their counterparts. Living in a slum area (AOR: 1.38; 95% CI: 1.06–1.79) was associated with food insecurity.

## DISCUSSION

This study assessed household food insecurity during the COVID-19 pandemic. Most household heads were aged at least 40 years (72%). The mean household size was six persons or fewer (75%), approximately one-third of the households were overcrowded (29%), and the SES distribution among households was equal. More than half the households were affected by food security conditions during the pandemic, with more than half frequently facing food insecurity. The prevalence of food insecurity was notably high during the pandemic, with significant disparities observed between non-slum and slum areas. Being aged 50 years or older, having an insufficient living space, having a certain SES (poorest, poorer, middle, or wealthier), and residing in a slum area were associated with a heightened risk of food insecurity.

These findings align with prior research demonstrating that economic shocks can lead to decreased food accessibility in vulnerable households; thus, the progression of the pandemic made it possible for food security to deteriorate further [2, 4, 6, 8–10, 20, 21].

Lockdown measures associated with the pandemic could have disrupted food supply chains and dietary habits, potentially contributing to various forms of malnutrition, including an increased risk of obesity owing to the consumption of highly processed foods and reduced physical activity [2]. Therefore, it is essential to closely monitor the indirect health effects of the COVID-19 pandemic.

Food insecurity was more prevalent in informal settlement households, in which more than three-quarters of the households reported having experienced this issue. Limited access to food resources exacerbates the adverse effects of the pandemic and obstructs infection control measures. Focused social protection actions are needed against shock, accounting for household profiles [2, 3, 8]. Vulnerable households, particularly those with informal jobs and young children, could benefit from cash transfer strategies to protect their food security. The expansion of food assistance programs, including cash transfers and food supply initiatives, should target overcrowded households and the informal job sector.

The findings related to the worsening trend of food insecurity emphasize the urgent need for policy actions addressing broader factors contributing to food insecurity, such as economic development, infrastructure, and governance. Providing and evaluating not only food assistance but also cash transfer initiatives, at least for the most vulnerable households, is important. A cash transfer strategy for households with informal jobs and children younger than five years may help protect their food security status. Monitoring food insecurity during shock events, particularly among older individuals and households with informal employment, is recommended [3, 8]. Addressing inequities in accessing physical and social infrastructure is crucial for food security beyond household income, as evidenced by Frayne and McCordic’s study in South Africa [22]. Households lacking consistent access to cooking fuel, medical care, electricity, or water, in addition to cash income, demonstrated significantly higher odds of being categorized as food insecure. These results imply that the conditions of an urban environment might better predict and explain food security than income alone. Similarly, Khan et al. highlighted the significance of resilience—such as infrastructure, women empowerment, economic performance, human capital, and emergency workforce—in mitigating the impact of crises, such as natural disasters, in low-income countries [23]. These studies emphasize the need to consider broader social and structural factors in strategies to enhance food security and reduce negative outcomes in large-scale crises.

Moreover, addressing interconnected syndemic-like crises warrants a multifaceted approach guided by data-driven strategies grounded in a flexible and intricate systems framework [5]. Importantly, even as the COVID-19 pandemic recedes, its far-reaching economic, health, and societal consequences are expected to persist over an extended period [11].

Notably, compared with non-slum households, slum households reported a significantly greater incidence rate of food insecurity. Food insecurity affected more than three-quarters of the slum households. This finding underscores that the COVID-19 pandemic has had adverse effects on food security across the socioeconomic spectrum, with particularly severe consequences for those with low SES. Thus, it is imperative to establish robust mechanisms for monitoring the food insecurity status of households during shock events, paying particular attention to older individuals and households that rely on informal employment. A study conducted in Mexico during the COVID-19 pandemic recommended monitoring for food insecurity in the general population, including critical vulnerable groups such as those with low- and middle-SES [2, 24, 25]. However, it is crucial to acknowledge the limitation of using the SES scale, which may not capture changes in economic circumstances resulting from the pandemic and would instead reflect only pre-pandemic SES.

This study had several strengths, including the comprehensive assessment of food insecurity on a large scale during the COVID-19 pandemic, which allows for generalizability across all of Kinshasa and other similar urban centers in the Democratic Republic of Congo and other sub-Saharan countries characterized by the presence of slums and non-slum areas. However, certain limitations must be acknowledged. First, this study did not assess the broader multisectoral impacts of COVID-19 beyond its influence on food security.

Additionally, the pre-pandemic food security conditions of households were not evaluated, thereby preventing direct comparisons of conditions before and during the pandemic.

Despite employing random sampling, we lack the means to confirm the representativeness of our sample in relation to the population of Kinshasa. The DRC last conducted a census in 1984, and there is no clear indication of whether our sample accurately represents the population of Kinshasa. Gender considerations, albeit rarely openly addressed in the DRC, could impact food security in Kinshasa. This study is also limited by the lack of collected information on gender. Future research should analyze how food insecurity and the factors contributing to it develop, particularly in response to the economic consequences of the COVID-19 pandemic. Social and structural determinants potentially contributing to food security, such as women’s empowerment and access to water, electricity, and medical care, should be further researched.

## CONCLUSION

The COVID-19 pandemic impacted household food security in Kinshasa and has been a primary contributor to heightened food insecurity conditions among households, potentially resulting in adverse consequences. Thus, governments must develop targeted strategies aimed at mitigating household vulnerability during periods of crisis. Additionally, to combat economic restrictions that lead to food insecurity during crises, policymakers and implementing partners might enhance food assistance programs, such as cash transfers and food supply initiatives, focusing on overcrowded households and the informal job sector.

## Data Availability

All the relevant data for this study are available from KSPH. The materials will be made available by the leading author upon request. Dataset can be found also in osf: https://osf.io/h4jt2/?view_only=ad40b39d0f584bf480fb04571ed86b70

## DECLARATIONS

### Ethics approval and consent to participate

Ethical clearance was obtained from the ethical review board of the Kinshasa School of Public Health (KSPH; no. ESP/CE/71B/2022). Written informed consent was obtained from each respondent during the data collection process. Privacy and confidentiality were maintained throughout this study.

### Consent for publication

Informed consent was obtained from all the participants involved in this study.

## Authors’ contributions

Conceptualization, APZ and MDK; methodology, APZ, TNT, BS, EL, KB, MBK, KF, KDM, and MDK; software, APZ, TNT, KF, and KDM; validation, APZ, TNT, BS, EL, KB, MBK, KF, KDM, and MDK; formal analysis, APZ, TNT, KF, KDM, and MDK; investigation, APZ, TNT, BS, EL, KB, MBK, KF, KDM, and MDK; resources, APZ, BS, EL, KB, MBK, KF, KDM, and MDK; data curation, APZ, KB, KF, KDM, and MDK; writing—original draft preparation: APZ, TNT, BS, EL, KB, MBK, KF, KDM, and MDK; writing—review and editing: APZ, TNT, BS, EL, KB, MBK, KF, KDM, and MDK; visualization, APZ, TNT, KF, KDM, and MDK; supervision, APZ, TNT, and MDK; project administration, APZ and MDK; and funding acquisition, APZ and MDK. All the authors have read and agreed to the final version of the manuscript. All authors consented to the publication of the most recent version of the present article.

## Acknowledgments

The authors thank the KSPH students who were involved in data collection. We also thank all respondents for sharing their experiences with us.

## LIST OF ABBREVIATIONS

AOR: Adjusted odds ratio
CI: Confidence interval
DRC: Democratic Republic of the Congo
EA: Enumeration area
HFIAS: Household Food Insecurity Access Scale
KSPH: Kinshasa School of Public Health
SES: Socioeconomic status

## Author information

Pierre Akilimali, the corresponding author, is a Professor at the University of Kinshasa School of Public Health.

